# Evaluation of the diagnostic value of YiDiXie^™^-SS, YiDiXie^™^-HS and YiDiXie^™^-D in gallbladder cancer

**DOI:** 10.1101/2024.09.18.24313930

**Authors:** Huimei Zhou, Pengwu Zhang, Chen Sun, Zhenjian Ge, Wenkang Chen, Yingqi Li, Shengjie Lin, Yutong Wu, Wuping Wang, Siwei Chen, Xutai Li, Wei Li, Yongqing Lai

## Abstract

**Background:** Gallbladder cancer is a grave threat to human health and poses a severe economic burden. Enhanced CT is extensively used in the diagnosis of gallbladder tumors. However, false-positive results on enhanced CT can lead to misdiagnosis and incorrect surgery or treatment, while false-negative results on enhanced CT can lead to missed diagnosis and delayed treatment. There is an urgency to find convenient, cost-effective and noninvasive diagnostic methods to decrease the false-positive and false-negative rates of gallbladder-enhanced CT. The goal of this study was to assess the diagnostic value of YiDiXie™-SS, YiDiXie™-HS and YiDiXie™-D in gallbladder cancer.

**Patients and methods:** Fifty study subjects (malignant group, n=12; benign group, n=38 cases) were finally recruited in the study. Remaining serum samples from the subjects were collected and tested by applying YiDiXie ™ all-cancer detection kit (YiDiXie™ all-cancer detection kit) to assess the sensitivity and specificity of YiDiXie™-SS, YiDiXie™-HS and YiDiXie™-D, respectively.

**Results:** The sensitivity of YiDiXie™ SS was 100% (95% CI: 75.8% - 100%) and its specificity was 65.8% (95% CI: 49.9% - 78.8%). This means that YiDiXie™ SS has very high sensitivity and high specificity in gallbladder tumors.The sensitivity of YiDiXie™-HS was 83.3% (95% CI: 55.2% - 97.0%) and its specificity was 84.2% (95% CI: 69.6% - 92.6%). This means that YiDiXie™-HS has high sensitivity and high specificity in gallbladder tumors.The sensitivity of YiDiXie™-D was 66.7% (95% CI: 39.1% - 86.2%) and its specificity was 92.1% (95% CI: 79.2% - 97.3%). This means that YiDiXie ™ -D has high sensitivity and very high specificity in gallbladder tumors. The sensitivity of YiDiXie™-SS in patients with positive enhanced CT was 100% (95% CI: 64.6% - 100%), and its specificity was 60.0% (95% CI: 31.3% - 83.2%). It implies that the application of YiDiXie™-SS reduces the false-positive rate of gallbladder-enhanced CT by 60.0% (95% CI: 31.3% - 83.2%) without essentially increasing the leakage of malignancies. The sensitivity of YiDiXie™-HS in enhanced CT-negative patients was 80.0% (95% CI: 37.6% - 99.0%) and its specificity was 85.7% (95% CI: 68.5% - 94.3%). It implies that the application of YiDiXie ™ -HS lowers the false-negative rate of enhanced CT by 80.0% (95% CI: 37.6% - 99.0%). The sensitivity of YiDiXie™-D in patients with positive enhanced CT was 71.4%(95% CI: 35.9% -94.9%) and its specificity was 90.0%(95% CI: 59.6% - 99.5%). It implies that YiDiXie™-SS lowers the false-positive rate of enhanced CT for 90.0%(95% CI: 59.6% - 99.5%). YiDiXie™-D has a sensitivity of 60.0% (95% CI: 23.1% - 92.9%) in patients with negative enhanced CT and its specificity is 92.9% (95% CI: 77.4% - 98.7%). This means that YiDiXie™-D reduces the false-negative rate of enhanced CT by 60.0% (95% CI: 23.1% - 92.9%) while maintaining high specificity.

**Conclusion:** YiDiXie ™ -SS has very high sensitivity and high specificity in gallbladder tumors.YiDiXie ™ -HS has high sensitivity and high specificity in gallbladder tumors.YiDiXie ™ -D has high sensitivity and very high specificity in gallbladder tumors.YiDiXie™ -SS significantly reduced gallbladder enhanced CT false-positive rates with essentially no increase in delayed treatment for gallbladder cancer. YiDiXie ™ -HS significantly reduces the false-negative rate of gallbladder enhanced CT. YiDiXie™ -D can significantly reduce the false-positive rate of gallbladder enhanced CT, or significantly reduce the false-negative rate of gallbladder enhanced CT while maintaining a high level of specificity.

**Clinical trial number:** ChiCTR2200066840.

## INTRODUCTION

Gallbladder cancer refers to malignant tumors that occur in the gallbladder (including the base, body, neck, and ducts of the gallbladder). Gallbladder cancer accounts for approximately 80% to 95% of biliary tract tumors and is the most common biliary tract malignancy^1-2^. The most recent data show an increasing trend in new cases and deaths with 115,949 new gallbladder cancer cases and 84,695 new deaths globally in 2020^3^.In 2022 there were 122,462 new gallbladder cancer cases and 89,031 new deaths globally^4^, with an increasing trend in new cases and deaths. In 2023 there were an estimated 12,350 diagnosed with gallbladder or other biliary tract cancers in the U.S., and approximately 4,530 deaths from gallbladder or other biliary tract cancers^5^. There are significant geographic, racial, and ethnic differences in its incidence, with women generally having more patients than men globally^6-7^. Geographically, gallbladder cancer is even the second leading cause of death in women after breast cancer in certain countries such as Chile. Similarly, India has a high burden of gallbladder cancer worldwide, with age-standardized incidence and mortality rates of 2.1/100,000 and 1.8/100,000 women, respectively^7-8^. Surgical resection is the only treatment with curative intent for gallbladder cancer, but very few cases are amenable to resection and most adjuvant treatments have very low response rates. This is mainly due to the fact that patients with gallbladder cancer are rarely symptomatic and are usually diagnosed at an advanced stage with a very poor prognosis. It is more difficult to detect gallbladder cancer early in the curable stage, and nearly 4/5 gallbladder cancer patients are diagnosed with advanced or metastatic stages, making gallbladder cancer one of the cancers with a very poor prognosis^9^. The 5-year survival rate of gallbladder cancer patients is only around 5%-19%^10-11^. Therefore, gallbladder cancer is a serious threat to human health. Therefore, gallbladder cancer is not only a serious threat to human health, but also a heavy burden to the patient, family, and society.

Enhanced CT is widely used in the diagnosis of gallbladder tumors. Enhanced CT scan can show the morphology of gallbladder cancer, the extent of gallbladder wall invasion, whether adjacent organs are involved, hemodynamic changes and lymph node metastasis, so as to evaluate the lesion^12-13^. However, the diagnostic level of CT is more affected by the level of physicians, external influences, different imaging features that are difficult to identify, and the accuracy of diagnosis is difficult to control^14-15^. On the one hand, enhanced CT can produce a large number of false-positive results. The rate of false-positive gallbladder-enhanced CT is about 12%-36%^16-18^. When enhanced CT is positive, patients usually undergo radical resection^19-20^. False-positive enhanced CT results mean that benign diseases are misdiagnosed as malignant tumors, and patients will have to bear the adverse consequences of unnecessary mental suffering, expensive surgery and examination costs, physical injuries, and even organ removal and loss of function. Therefore, there is an urgent need to find a convenient, cost-effective, and noninvasive diagnostic method to reduce the false-positive rate of gallbladder-enhanced CT.

On the other hand, enhanced CT can produce a large number of false-negative results. One study showed that the rate of false-negative gallbladder-enhanced CT was 10.5%-45.7%^17-18,21^. When enhanced CT is negative, patients are usually taken for observation and regular follow-up^19-20^. False-negative enhanced CT results imply misdiagnosis of malignant tumors as benign disease, which will likely lead to delayed treatment, progression of malignant tumors, and possibly even development of advanced stages. Patients will thus have to bear the adverse consequences of poor prognosis, expensive treatment, poor quality of life, and short survival. False-negative results on enhanced CT also increase the risk of multiple follow-up examinations in the program itself, such as the adverse sequelae of contrast injection and the effects of radiation^22-23^. Therefore, there is an urgent need to find a convenient, cost-effective and noninvasive diagnostic method to reduce the false negative rate of gallbladder-enhanced CT.

In addition, there are some special patients who require extra caution in choosing whether or not to operate, such as smaller tumors and patients in poor general condition. The risk of wrong surgery in these special patients is much higher than the risk of missed diagnosis. False-positive results of enhanced CT can lead to misdiagnosis and wrong surgery or treatment, while false-negative results of enhanced CT can lead to missed diagnosis and delayed treatment. Therefore, there is an urgent need to find a convenient, cost-effective and noninvasive diagnostic method with high specificity to substantially reduce the false-positive rate of gallbladder enhanced CT in these special patients or to significantly reduce its false-negative rate while maintaining high specificity.

On the basis of detecting novel tumor markers of miRNA in serum, an in vitro diagnostic test product-YiDiXie ™ all-cancer test (hereinafter referred to as The YiDiXie ™ test) has been developed by Shenzhen KeRuiDa Health Science and Technology Co^24^. The YiDiXie ™ test The YiDiXie™ test detects a wide range of cancer types with only 200 microliters of whole blood or 100 microliters of serum at a time^24^. The YiDiXie™ test comprises three products with very distinct properties, YiDiXie™-HS, YiDiXie™-SS and YiDiXie™-D^24^.

The goal of this research is to assess the diagnostic value of YiDiXie™-SS, YiDiXie™-HS and YiDiXie™-D in gallbladder cancer.

## PATIENTS AND METHODS

### Study design

The current work is part of the sub-study “Assessment of the adjuvant diagnostic value of The YiDiXie ™ test in multiple tumors” of the SZ-PILOT study (ChiCTR2200066840).

The SZ-PILOT study (ChiCTR2200066840) is a prospective, single-center, observational study. Subjects who signed a pan-informed consent form for donation of residual samples at the time of admission or physical examination were enrolled, and 0.5 ml of their serum residual samples were obtained for the study.

The study was carried out in a blinded manner. Neither the laboratory worker who performed The YiDiXie ™ test nor the KeRuiDa laboratory technician who determined the results of The YiDiXie™ test were aware of the subject’s clinical profile. The clinical experts who assessed the subjects’ clinical data were also unaware of the results of The YiDiXie™ test.

The study was approved by the Ethics Committee of Shenzhen Hospital of Peking University and was conducted in compliance with the International Conference on Harmonization (ICH) Code of Practice for the Quality Management of Pharmaceutical Clinical Trials and the Declaration of Helsinki.

### Participants

Subjects in both groups were enrolled separately, and all subjects who met the inclusion criteria were consecutively included.

Initially, inpatients with “suspected (solid or hematologic) malignancy” who signed a general informed consent for donation of the remaining samples were enrolled in this study. Postoperative pathological diagnosis of “malignant tumor” was included in the malignant tumor group, and post-operative pathology diagnosis of “benign disease” was incorporated into the benign disease group. The subjects with ambiguous pathologic results were excluded from the study. Part of the samples from the malignant tumor group were used in our preliminary work^24^.

Participants who did not pass the serum sample quality test prior to The YiDiXie™ test were not included in this study. Please refer to our preliminary work^24^ for specific enrollment and exclusion.

### Sample collection, processing

In this study, serum samples used were obtained from serum left over after normal treatment and no additional blood draws were required. About 0.5 ml of serum samples were obtained from the remaining serum of subjects in the Medical Laboratory Department and stored at -80°C for use in subsequent the YiDiXie™ test.

### The YiDiXie test

The YiDiXie™ test was done by using YiDiXie™ all-cancer detection kit. It is an in vitro diagnostic kit developed and produced by Shenzhen KeRuiDa Health Technology Company Limited for use with the fluorescent quantitative PCR instrument. For details, please see our preliminary work^24^.

The YiDiXie™ test contains 3 assays with quite distinct properties: YiDiXie™-HS, YiDiXie™-SS, and YiDiXie™-D. For a detailed description, please refer to our previous work^24^.

Conducting the YiDiXie™ test according to the instructions of the YiDiXie™ all-cancer detection kit. For detailed instructions, please refer to our previous work^24^.

The original test result was evaluated and analyzed by the technical staff of Shenzhen KeRuiDa Health Technology Company Limited, and the result of The YiDiXie™ test was determined to be either “positive” or “negative”^24^.

### Diagnosis of enhanced CT

“Positive” or “negative” results are determined based on the diagnostic findings of enhanced CT “Positive” is determined if the diagnosis is positive, more certain, or favors malignant tumors. “Negative” is determined if the diagnostic conclusion is positive, more certain, or favors benign disease, or if the diagnostic conclusion is ambiguous as to whether the disease is benign or malignant.

### Extraction of clinical data

All the clinical, pathological, laboratory and imaging data in the study were retrieved from the subjects’ inpatient medical records or physical examination reports. Classical staging was assessed and completed by trained clinicians in accordance with the AJCC staging manual (7th or 8th edition)^25-26^.

### Statistical analyses

Data for descriptive statistics were reported for demographic and baseline characteristics. The number and percentage of subjects in each category were computed for categorical variables. It was calculated for continuous variables the total number of subjects (n), mean, standard deviation (SD) or standard error (SE), median, first quartile (Q1), third quartile (Q3), minimum and maximum values. Calculation of 95% confidence intervals (CI) for multiple indicators was done with Wilson (score) method.

## RESULTS

### Participant disposition

A total of 50 study participants (malignant group, n=12; benign group, n=38) were eventually included in this study. In Table 1, we listed the demographic and clinical characteristics of the 50 study participants.

**Table 1.** Participants’ demographic and clinical manifestation

|  | Malignant (n = 12) | Benign (n = 38) | Total (N = 50) |
| --- | --- | --- | --- |
| Age, years |  |  |  |
| Mean (SD) | 63.6 (7.92) | 44.5 (13.06) | 49.1 (14.51) |
| Median (Q1,Q3) | 65 (58, 68) | 44 (33, 52) | 49 (40, 58) |
| Min, max | 46, 76 | 22, 79 | 22, 79 |
| Age, group, n (%) |  |  |  |
| < 50 | 1 (8.3) | 26 (68.4) | 27 (54.0) |
| ≥ 50 | 11 (91.7) | 12 (31.6) | 23 (46.0) |
| < 65 | 5 (41.7) | 35 (92.1) | 40 (80.0) |
| ≥ 65 | 7 (58.3) | 3 (7.9) | 10 (20.0) |
| Sex, n (%) |  |  |  |
| Female | 9 (75.0) | 15 (39.5) | 24 (48.0) |
| Male | 3 (25.0) | 23 (60.5) | 26 (52.0) |
| Body mass index (kg/m2) |  |  |  |
| n | 12 | 37 | 49 |
| Mean (SD) | 24.3 (3.53) | 22.9 (2.97) | 23.2 (3.14) |
| Median (Q1,Q3) | 24.6 (22.6, 26.4) | 23.2 (20.5, 25.4) | 23.2 (21.2, 25.4) |
| Min, max | 15.6 28.9 | 17.1 29.2 | 15.6 29.2 |
| Body mass index category, n (%) |  |  |  |
| Underweight | 1 (8.3) | 2 (5.3) | 3 (6.0) |
| Benign | 4 (33.3) | 20 (52.6) | 24 (48.0) |
| Overweight | 6 (50.0) | 13 (34.2) | 19 (38.0) |
| Obese | 1 (8.3) | 2 (5.3) | 3 (6.0) |
| Missing | 0 (0) | 1 (2.6) | 1 (2.0) |
| AJCC clinical stage |  |  |  |
| Stage I | 2 (16.7) |  | 2 (16.7) |
| Stage II | 5 (41.7) |  | 5 (41.7) |
| Stage III | 0 (0) |  | 0 (0) |
| Stage IV | 4 (33.3) |  | 4 (33.3) |
| Missing | 1 (8.3) |  | 1 (8.3) |

Q1,Q3, first quartile, third quartile; SD, standard deviation.

Both groups of study participants were compared in terms of demographic and clinical characteristics (Table 1). Average (standard deviation) age was 49.1 (14.51) years and 48.0% (24/50) were female.

### Diagnostic performance of YiDiXie™-SS

As shown in Table 2, the sensitivity of YiDiXie™ SS was 100% (95% CI: 75.8% - 100%) and its specificity was 65.8% (95% CI: 49.9% - 78.8%). This means that YiDiXie ™ SS has very high sensitivity and high specificity in gallbladder tumors.

**Table 2.** The performance of YiDiXie™-SS

|  | Malignant | Benign | Total |
| --- | --- | --- | --- |
|  | 12 | 38 | 50 |
| Positive | 12 | 13 | 25 |
| Negative | 0 | 25 | 25 |

SEN = $12/12 = 100\%$ (75.8% - 100%) SPE = $25/38 = 65.8\%$ (49.9% - 78.8%)
SEN, Sensitivity. SPE, Specificity. Two-sided 95% Wilson confidence intervals were calculated.

### Diagnostic performance of YiDiXie™-HS

As shown in Table 3, the sensitivity of YiDiXie™ -HS was 83.3% (95% CI: 55.2% - 97.0%) and its specificity was 84.2% (95% CI: 69.6% - 92.6%). This means that YiDiXie ™ -HS has high sensitivity and high specificity in gallbladder tumors.

**Table 3.** The performance of YiDiXie™-HS

|  | Malignant | Benign | Total |
| --- | --- | --- | --- |
|  | 12 | 38 | 50 |
| Positive | 10 | 6 | 16 |
| Negative | 2 | 32 | 34 |

SEN = $10/12 = 83.3\%$ (55.2% - 97.0%) SPE = $32/38 = 84.2\%$ (69.6% - 92.6%)
SEN, Sensitivity. SPE, Specificity. Two-sided 95% Wilson confidence intervals were calculated.

### Diagnostic performance of YiDiXie™-D

As shown in Table 4, the sensitivity of YiDiXie™ -D was 66.7% (95% CI: 39.1% - 86.2%) and its specificity was 92.1% (95% CI: 79.2% - 97.3%). This means that YiDiXie ™ -D has high sensitivity and very high specificity in gallbladder tumors.

**Table 4.** The performance of YiDiXie™-D

|  | Malignant | Benign | Total |
| --- | --- | --- | --- |
|  | 12 | 38 | 50 |
| Positive | 8 | 3 | 11 |
| Negative | 4 | 35 | 39 |

SEN, Sensitivity. SPE, Specificity. Two-sided 95% Wilson confidence intervals were calculated.

### Diagnostic performance of enhanced CT

As shown in Table 5, the sensitivity of enhanced CT was 58.3% (95% CI: 32.0% - 80.7%) and its specificity was 73.7% (95% CI: 58.0% - 85.0%).

**Table 5.** Performance of different Items

| Items | Total N | Positive | Sensitivity % (95% CI)<sup>a</sup> | Total N | Negative | Specificity % (95% CI)<sup>b</sup> |
| --- | --- | --- | --- | --- | --- | --- |
| CT | 12 | 7 | 58.3% (32.0% - 80.7%) | 38 | 28 | 73.7% (58.0% - 85.0%) |
| YiDiXie™-SS in CT-positive | 7 | 7 | 100% (64.6% - 100%) | 10 | 6 | 60.0% (31.3% - 83.2%) |
| YiDiXie™-HS in CT-negative | 5 | 4 | 80.0% (37.6% - 99.0%) | 28 | 24 | 85.7% (68.5% - 94.3%) |
| YiDiXie™-D in CT-positive | 7 | 5 | 71.4% (35.9% - 94.9%) | 10 | 9 | 90.0% (59.6% - 99.5%) |
| YiDiXie™-D in CT-negative | 5 | 3 | 60.0% (23.1% - 92.9%) | 28 | 26 | 92.9% (77.4% - 98.7%) |

CI, confidence interval. <sup>a,b</sup> Two-sided 95% Wilson confidence intervals were calculated.

### Diagnostic Performance of YiDiXie™-SS in enhanced CT-positive patients

To address the challenge of high false-positive rate of gallbladder enhanced CT, YiDiXie™-SS was applied to enhanced CT-positive patients.

The sensitivity of YiDiXie™-SS in patients with positive enhanced CT was 100% (95% CI: 64.6% - 100%), and its specificity was 60.0% (95% CI: 31.3% - 83.2%), as shown in Table 5. It implies that the application of YiDiXie ™ -SS reduces the false-positive rate of gallbladder-enhanced CT by 60.0% (95% CI: 31.3% - 83.2%) without essentially increasing the leakage of malignancies.

### Diagnostic Performance of YiDiXie™-HS in enhanced CT-negative patients

To address the challenge of high false-negative rate of gallbladder enhanced CT, YiDiXie™-HS was applied to enhanced CT-negative patients.

The sensitivity of YiDiXie ™ -HS in enhanced CT-negative patients was 80.0% (95% CI: 37.6% - 99.0%) and its specificity was 85.7% (95% CI: 68.5% - 94.3%), as shown in Table 5. It implies that the application of YiDiXie ™ -HS lowers the false-negative rate of enhanced CT by 80.0% (95% CI: 37.6% - 99.0%).

### Diagnostic performance of YiDiXie™-D in enhanced CT-positive patients

The false-positive consequences are significantly worse than the false-negative consequences in certain patients with positive enhanced CT. Therefore, applying YiDiXie ™ -D to such patients to reduce their false-positive rate.

The sensitivity of YiDiXie™-D in patients with positive enhanced CT was 71.4%(95% CI: 35.9% -94.9%) and its specificity was 90.0%(95% CI: 59.6% - 99.5%), as shown in Table 5. It implies that YiDiXie™ -SS lowers the false-positive rate of enhanced CT for 90.0%(95% CI: 59.6% - 99.5%).

### Performance of YiDiXie™-D in patients with enhanced CT negative results

The false-positive consequences are significantly worse than the false-negative consequences in certain patients with negative enhanced CT, so the more specific YiDiXie ™ -D should be applied to such patients.

As shown in Table 5, YiDiXie ™ -D has a sensitivity of 60.0% (95% CI: 23.1% - 92.9%) in patients with negative enhanced CT and its specificity is 92.9% (95% CI: 77.4% - 98.7%). This means that YiDiXie™-D reduces the false-negative rate of enhanced CT by 60.0% (95% CI: 23.1% - 92.9%) while maintaining high specificity.

## DISCUSSION

### Clinical significance of YiDiXie™-SS in enhanced CT-positive patients

Further diagnostic methods are significant for both sensitivity and specificity in patients with positive enhanced CT of the gallbladder. A higher false-negative rate implies that more malignancies are underdiagnosed, which will cause a delay in their treatment, a progression of the malignancy, and possibly even the progression to an advanced stage. More false-positive rates indicate more misdiagnosis of benign disease, which may lead to unnecessary surgery. Generally, when benign gallbladder disease is misdiagnosed as a malignant tumor, it usually will likely cause unnecessary surgery, which will not affect the patient’s prognosis, and its cost of treatment is much lower than that of late-stage cancer.

Furthermore, in patients with positive gallbladder-enhanced CT, the positive predictive value is higher. Even if the false-negative rate is comparable to the false-positive rate, it is still more harmful. Hence, a very high sensitivity but slightly lower specificity YiDiXie ™ -SS was chosen for reducing the gallbladder-enhanced CT false-positive rate.

Sensitivity of YiDiXie ™ -SS in patients with positive enhanced CT was 100% (95% CI: 64.6% - 100%) and its specificity was 60.0% (95% CI: 31.3% - 83.2%), as shown in Table 5. Based on the above results, it was shown that YiDiXie™-SS could reduce the false-positive rate of gallbladder enhanced CT by 60.0% (95% CI: 31.3% - 83.2%), keeping the sensitivity approaching 100%.

It implies that YiDiXie™-SS dramatically lowers the probability of misdiagnosis of benign gallbladder disease as a malignant tumor with essentially no increase in malignancy delayed treatment. Hence, YiDiXie ™ -SS fulfills the clinical demand well and is clinically important in gallbladder cancer.

### Clinical significance of YiDiXie™-HS in enhanced CT-negative patients

Further diagnostic methods are important for both sensitivity and specificity in patients with negative enhanced CT of the gallbladder. More false-negative rates imply that more gallbladder cancers are underdiagnosed, which would result in a delay in their treatment, malignant progression, and possibly even advanced stages. Generally, patients with benign gallbladder disease misdiagnosed as malignant tumors usually undergo surgery, it does not affect the patient’s prognosis, and the costs of its treatment are much lower compared with that of advanced cancers. Thus, for patients with negative enhanced CT, the “risk of missed diagnosis of gallbladder cancer” is higher than the “risk of misdiagnosis of benign gallbladder disease”.

Additionally, a higher negative predictive value was observed in patients with negative gallbladder-enhanced CT. Both higher false-positive and false-negative rates can lead to significant harm. Hence, the high sensitivity and specificity of YiDiXie ™ -HS was chosen for the reduction of gallbladder-enhanced CT false-negative rate.

The sensitivity of YiDiXie™-HS was 80.0% (95% CI: 37.6% - 99.0%) and its specificity was 85.7% (95% CI: 68.5% - 94.3%) in patients with negative enhanced CT, as shown in Table 5. Above results showed that the utilization of YiDiXie ™ -HS decreased the false-negative rate of enhanced CT by 80.0% (95% CI: 37.6% - 99.0%).

It implies that YiDiXie™-HS greatly decreases the possibility of a negative enhanced CT for malignant tumor diagnosis. As a result, YiDiXie ™ -HS well fulfills the clinical needs and is clinically significant in gallbladder cancer.

### Diagnostic performance of YiDiXie™-D

Gallbladder tumors considered malignant usually undergo radical resection. In some cases, however, extra caution is needed in selecting whether or not to operate. Hence further diagnosis is required, for example: smaller tumors, poor general condition of the patient, etc.

The sensitivity and specificity of further diagnostic methods are crucial in patients with gallbladder tumors. Weighing the conflict between sensitivity and specificity is essentially a trade-off between the “risk of underdiagnosis of gallbladder cancer” and the “risk of misdiagnosis of benign gallbladder disease”. As small tumors have a lower risk of tumor progression and distant metastasis, the risk of underdiagnosis of gallbladder cancer is much lower than the risk of misdiagnosis of benign gallbladder diseases. The risk of misdiagnosis of benign gallbladder disease is much higher than the risk of misdiagnosis of gallbladder cancer in patients with poor general conditions because the perioperative risk is much higher than the general conditions. Therefore, for these patients, YiDiXie™ -D, which has a very high specificity but a slightly lower sensitivity, was chosen to reduce the rate of false positives on gallbladder enhanced CT or to significantly reduce the rate of false negatives while maintaining a high specificity.

As shown in Table 5, the sensitivity of YiDiXie™ -D in patients with positive enhanced CT was 71.4% (95% CI: 35.9% - 94.9%), and its specificity was 90.0% (95% CI: 59.6% - 99.5%); the sensitivity of YiDiXie™-D in patients with negative enhanced CT was 60.0% (95% CI: 23.1% - 92.9%), and its specificity was 92.9% (95% CI: 77.4% - 98.7%). These results suggest that YiDiXie ™ -D can significantly reduce the false-positive rate of gallbladder enhanced CT, or significantly reduce the false-negative rate of gallbladder enhanced CT while maintaining high specificity.

It implies that YiDiXie ™ -D greatly decreases the possibility of incorrect surgery in these patients with gallbladder tumors that demand extra cautious surgery. Thus, it is clear that YiDiXie™-D is well suited to fulfill the clinical needs and is of great clinical importance in gallbladder cancer.

### The YiDiXie™ test are expected to solve 2 challenges in brain tumors

The first is that YiDiXie™-SS markedly reduces the risk of misdiagnosis of benign gallbladder disease as gallbladder cancer. From one side, YiDiXie ™ -SS substantially reduces the probability of incorrect surgery for benign gallbladder disease while essentially not increasing the underdiagnosis of gallbladder cancer. With essentially no increase in gallbladder cancer miss-diagnosis, YiDiXie™-SS reduced the rate of false-positive gallbladder enhanced CT by 60.0% (95% CI: 31.3% - 83.2%), as shown in Table 5. Thus, YiDiXie ™ -SS markedly reduces the range of adverse outcomes associated with unnecessary surgery without essentially increasing delayed treatment of gallbladder cancer.

On the other hand, YiDiXie ™ -SS provides significant relief to surgeons from non-essential work. In the case of a positive enhanced CT of the gallbladder, the patient usually undergoes radical surgery. The ability to perform these radical surgeries in a timely manner is directly dependent on the number of surgeons. Appointments are available for months or even more than a year in many areas of the world. Unavoidably, this delays the treatment of gallbladder cancer among them, and consequently it is not uncommon for gallbladder enhanced CT-positive patients awaiting radical surgery to have progression of malignancy or even distant metastases. YiDiXie ™ -SS could reduce the false-positive rate of gallbladder-enhanced CT by 60.0% (95% CI: 31.3% - 83.2%) with essentially no increase in missed diagnosis of gallbladder cancer, as shown in Table 6. Hence, YiDiXie ™ -SS greatly relieves surgeons from unnecessary workload and facilitates timely diagnosis and treatment of gallbladder cancer or other conditions for which treatment would have been delayed.

Furthermore, YiDiXie™-HS greatly reduces and minimizes the risk of underdiagnosis of gallbladder cancer. A negative enhanced CT usually rules out gallbladder cancer for the time being. Because of the high rate of false-negative enhanced CT, it causes massive delays in the treatment of gallbladder cancer patients. As shown in Table 7, YiDiXie™-HS decreased the false negative rate of enhanced CT by 80.0% (95% CI: 37.6% - 99.0%). Thus, YiDiXie ™ -HS greatly decreased the probability of missed diagnosis of gallbladder cancer on enhanced CT, promoting timely diagnosis and treatment of patients with gallbladder cancer who were otherwise delayed in treatment.

Again, YiDiXie ™ -D is expected to solve the problem of “ high false-positive rate “ and “ high false-negative rate “ in some specific patients. Gallbladder tumors are usually operated on when malignancy is considered. However, on smaller tumors, poor general condition of the patient, etc., surgery may result in unnecessary surgical trauma, organ removal, loss of function, or the development of serious perioperative complications. Hence there is a need for extra caution in these patients before surgery. As shown in Table 5, YiDiXie™-SS reduced the false-positive rate of enhanced CT by 90.0% (95% CI: 59.6% - 99.5%) or, while maintaining a high level of specificity, reduced the false-negative rate of enhanced CT by 60.0% (95% CI: 23.1% - 92.9%). Thus, YiDiXie ™ -D sharply lowered the rate of risk of wrongful surgery or serious perioperative complications that occurred during surgery with these patients.

Final, the YiDiXie ™ test allows for a “just-in-time” diagnosis of gallbladder tumors. On the one hand, the YiDiXie™ test needs only a small amount of blood, allowing patients to complete the diagnostic process non-invasively in the comfort of their own home. only 0.2 ml of full blood is sufficient to conduct the YiDiXie ™ test^24^. The general population can obtain 0.2 mL of finger blood at home using a finger blood collection needle, instead of requiring a medical staff to take blood from a vein^24^. Thus, the diagnostic process can be accomplished non-invasively without the patient having to leave their home.

In addition, there is almost unlimited diagnostic capacity in the YiDiXie™ test. The basic flowchart of the YiDiXie™ test is shown in Figure 1, which shows that the YiDiXie ™ test needs no doctor or medical device, as well as no medical personnel to collect blood. Consequently, the number of medical personnel and medical facilities is completely independent of the YiDiXie ™ test, and its detection capacity is virtually unlimited. This enables the YiDiXie ™ test to “just-in-time” diagnose gallbladder tumors with no anxiety about waiting for an appointment.

**Figure 1.**
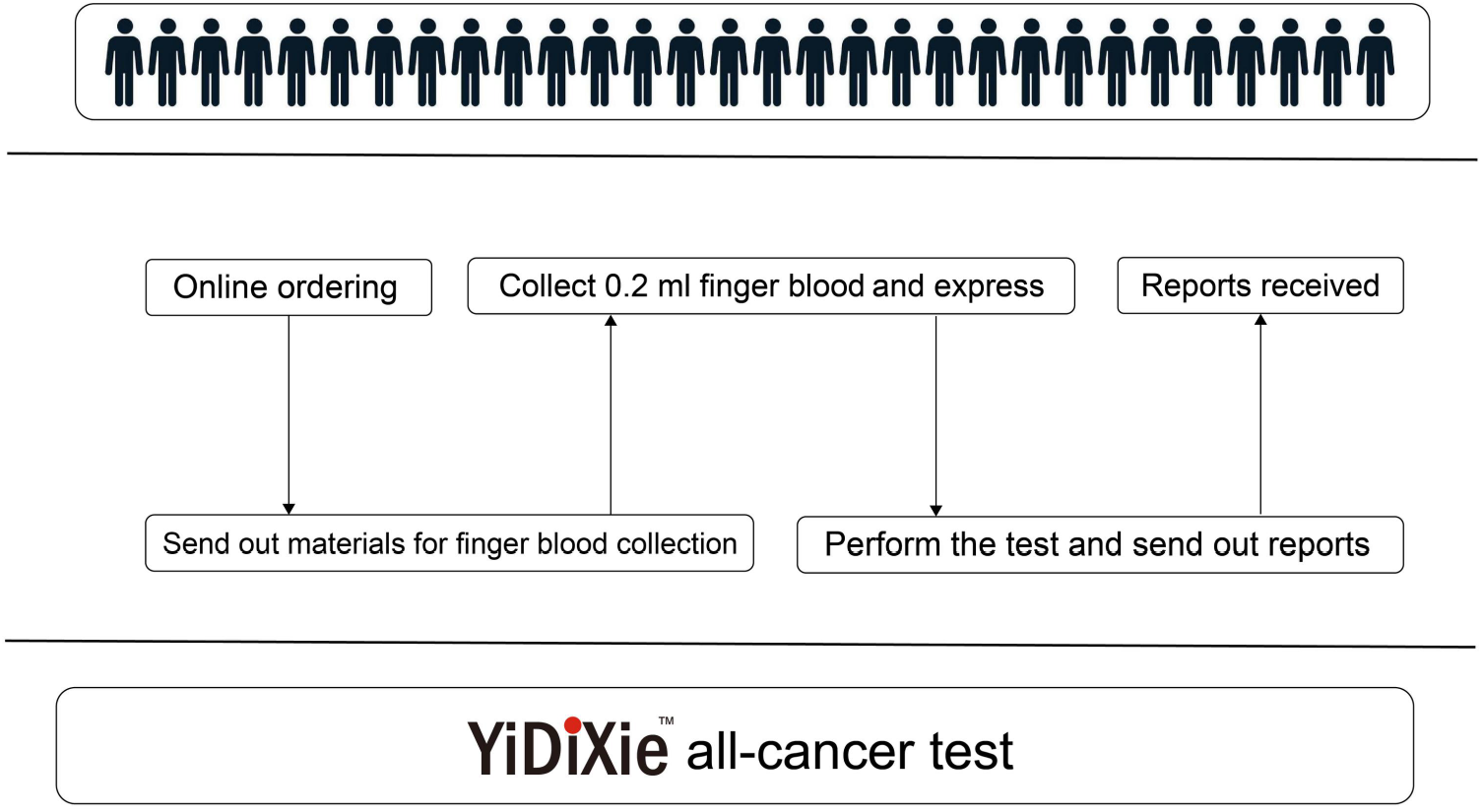
Basic flowchart of the YiDiXie™ test.

Briefly, the YiDiXie ™ test provides critical diagnostic value in gallbladder tumors, potentially solving the problems of “high false-positive rate of enhanced CT” and “high false-negative rate of enhanced CT” in gallbladder tumors.

### Limitations of the study

Firstly, the case number in this study was small and requires future clinical studies with larger sample sizes for further assessment.

Next, the present study included hospitalized patients with gallbladder cancer and benign gallbladder disease, which needs a cohort study of the natural population of gallbladder tumors for further evaluation in the future.

Lastly, our study was a single-center study, which may lead to a certain degree of bias in the results of this study. A multicenter study is needed to further evaluate this in the future.

## CONCLUSION

YiDiXie™-SS has very high sensitivity and high specificity in gallbladder tumors.YiDiXie ™ -HS has high sensitivity and high specificity in gallbladder tumors.YiDiXie ™ -D has high sensitivity and very high specificity in gallbladder tumors.YiDiXie™-SS significantly reduced gallbladder enhanced CT false-positive rates with essentially no increase in delayed treatment for gallbladder cancer. YiDiXie™ -HS significantly reduces the false-negative rate of gallbladder enhanced CT. YiDiXie ™ -D can significantly reduce the false-positive rate of gallbladder enhanced CT, or significantly reduce the false-negative rate of gallbladder enhanced CT while maintaining a high level of specificity.

## Data Availability

All data produced in the present study are contained in the manuscript.

